# Motor and Nonmotor Features of p.A53T Alpha-Synuclein PD vs idiopathic PD: Longitudinal Data from the PPMI Study

**DOI:** 10.1101/2024.08.23.24312442

**Authors:** Athina Maria Simitsi, Evangelos Sfikas, Christos Koros, Nikolaos Papagiannakis, Ion Beratis, Dimitra Papadimitriou, Roubina Antonellou, Stella Fragiadaki, Dionysia Kontaxopoulou, Marina Picillo, Ioanna Pachi, Ioanna Alefanti, Maria Stamelou, Paolo Barone, Leonidas Stefanis

## Abstract

**Background and Objectives:** The phenotype of p.A53T-Parkinson’s Disease (PD) mutation carriers appears more severe than that of iPD patients, however information on comparative, prospective clinical evolution of such subjects is limited. Here we conducted a longitudinal study to investigate, using multiple parameters, the progression of motor and nonmotor features of p.A53T SNCA PD compared to iPD.

**Methods:** Longitudinal 3-year data, concerning both motor and non-motor features, of 16 p.A53T-PD and 48 iPD, matched for age and disease duration at baseline, were downloaded from the Parkinson’s Progression Markers Initiative (PPMI) database and compared between the two groups. Additionally, a cognitive composite score was generated by 5 cognitive tests, focused more on executive/visuospatial function [Composite Score= (MOCA+ LNST+ SDMT+ BENTON+ Semantic F)/5], to better study cognitive change over a 3-year follow-up.

**Results:** Neuropsychological assessments (MOCA, LNST, BENTON, SDMT, Semantic fluency, Phonemic Fluency, HVLT.IM-REC, HVLT-RDLY) revealed significant cognitive decline in A53T-PD compared to iPD, but also across time for the group of A53T-PD. Especially in composite score, the group of A53T-PD had lower values at all time points vs baseline (p=0.004, p<0.001 and p<0.001 respectively), and vs iPD (p-values<0.001, at all time points, including baseline). Autonomic dysfunction using SCOPA-AUT, as well as specific items of MDS-UPDRS I, such as I.1. and I.2, reflecting cognitive function and psychotic features respectively, were more prominent both within the A53T-PD group over time and across several time points between the two groups. As far as motor symptoms are concerned, motor assessments, such as MDS-UPDRS.III.ON, H&Y. ON, S&E and MDS-UPDRS II showed worse performance in A53T-PD compared to iPD but also across time for the group of A53T-PD, even though LEDD was significantly higher in A53T-PD.

**Discussion:** The current longitudinal study indicates that A53T-PD represents a rapidly progressing subtype of PD, with accelerated decline in both motor and non-motor parameters, especially in cognitive function. Such data may set the stage for the application of targeted disease-modifying therapies in this particular subtype, in which the etiological link to alpha-synuclein is established, while generated data may be widely applicable to iPD, which is largely a sporadic synucleinopathy.

## 1. INTRODUCTION

The p.A53T mutation in the *SNCA* gene, encoding for the presynaptic protein alpha-synuclein (AS), was the first genetic alteration linked to Parkinson’s disease (PD). It was originally identified in families of Greek or Italian origin^1^. Other missense point mutations, as well as duplications or triplications of the *SNCA* gene, are also associated with familial autosomal dominant PD^2,3^. The AS protein is the principal component of Lewy bodies (LBs), the neuropathological hallmark of the disease in genetic and idiopathic PD (iPD). Patients with *SNCA* mutations are under intense scrutiny, given the manifest link of *SNCA* and AS to the pathophysiology, not only of familial, but also of iPD.

The phenotype of p.A53T mutation carriers is quite variable, but it is overall more aggressive than iPD^4,5^. The average onset age of A53T-related PD (A53T-PD) is around 46 years. The disease is highly but not completely penetrant (80%–90%)^6^. Non-motor features are prominent^6^, with hyposmia being more common^7^, and RBD occurring at a higher percentage compared to iPD^8^. The cognitive status is variable, ranging from intact to PD dementia (PDD), depending largely on disease duration^6^; frontal-executive and visuospatial functions are disproportionately affected compared to iPD^7^.

Longitudinal studies of motor and non-motor features in A53T-PD are scarce; the only previous study, performed by our team, used a rather basic assessment at two time points within 2 years and lacked a comparative control group^6^.

The Parkinson’s Disease Progression Marker Initiative (PPMI) database, with its extensive genetic information and comprehensive assessments, provides an excellent resource for investigating clinical, imaging and biochemical parameters in a large cohort of PD participants and in healthy controls.

In this study we investigated the longitudinal pattern of clinical manifestations in the p.A53T PPMI cohort compared to iPD. To this end, detailed longitudinal 3-year data, concerning both motor and non-motor features, were deployed from the PPMI study database for the two groups. This study provides novel insights into the disease trajectory of pA53T-carriers, pinpointing the main progressive features of this rare genetic cohort.

## 2. METHODS

### Data acquisition

This is a retrospective observational cohort study and the data used in the preparation of this article were obtained on October 05, 2023 from the PPMI database (https://www.ppmi-info.org/access-data-specimens/download-data), RRID:SCR_006431. For up-to-date information on the study, visit http://www.ppmi-info.org. This analysis used data openly available from PPMI. The present study was conducted in agreement with the principles of the Declaration of Helsinki and was approved by the Scientific Board of all PPMI sites involved. Signed informed consent was obtained from all participants recruited.

Demographic characteristics assessed were Age at BL visit, sex, years of education and PD duration. For each A53T-PD, we used propensity-based matching to identify 3 iPD as controls (1:3 ratio), ensuring these individuals had completed 3-year follow up visits in PPMI, according to the protocol described in the next section. Sex matching was not performed because the resulted original matching was statistically balanced between samples’ sex and avoided introducing further constraints. We reviewed data from 724 enrolled iPD, which was the original input control sample assessed for matching, to find the final control sample, consisting of 48 control iPD individuals, matched with 16 A53T-PD for age and PD duration at BL visit.

### Participant selection

In general, A53T-PD have younger onset of disease^6^, and the PPMI inclusion criteria do not define a maximum disease duration for them, in contrast to iPD (2 years). So, to avoid possible confounding effects from age and disease duration the selection process described below was followed.

Initially, a logistic regression model was fitted with the presence of A53T mutation as the dependent variable and age and disease duration as the independent variables. In the initial model all iPD visits that had at least 3 years of follow-up were included, but only the baseline visits of A53T-PD. Then we used the resulting model to compute a probability score for each subject-visit pair and using nearest neighbor matching with the score as distance.

In cases where a participant gave more than one matches, the nearest one was kept, and the process was rerun from the start with the worst pair removed from consideration. The algorithm was repeated until each iPD participant had only one matched visit.

### Clinical data

Longitudinal 3-year clinical data included assessments of motor symptoms, non-motor symptoms, and neurocognitive tests. These tools included the Movement Disorder Society Unified Parkinson’s Disease Rating Scale (MDS-UPDRS) Parts I-IV, Hoehn and Yahr scale (H&Y ON & OFF) Letter Number Sequencing (LNST), Hopkins Verbal Learning Test, Benton Judgment of Line Orientation Test (BENTON), Semantic Fluency (Semantic.F) & Phonemic Fluency Tests (Phonemic F), Symbol Digit Modalities Test (SDMT), Montreal Cognitive Assessment (MOCA), Geriatric Depression Scale (GDS), Questionnaire for Impulsive-Compulsive Disorders in Parkinson’s Disease (QUIP), Scales for Outcomes in Parkinson’s Disease-Autonomic (SCOPA-AUT), Epworth Sleepiness Scale (ESS), Rapid Eye Movement Sleep Behavior Disorder Screening Questionnaire (RBDQS), Activities of Daily Living (ADL) and L-dopa equivalent daily dose (LEDD). Additionally, a composite cognitive score was generated by 5 cognitive tests, focused more on executive/visuospatial function, Composite Score: (MOCA+ Semantic.F+ LNST+ BENTON+ SDMT)/5 (MOCA: global cognitive function, Semantic.F, LNST, SDMT: executive function, BENTON: visuospatial function).

### Statistical analysis

Means and standard deviations were used to describe the values of all markers examined. For the baseline comparisons for non-normally distributed scores, nonparametric Mann-Whitney U test was used. For dichotomous data, analysis was performed using Chi–square test. The effects of MOCA, BENTON, HVLT-RDLY, HVLT-REC, HVLT.IM-REC, LNST, Phonemic-F, SDMT, Semantic-F and Composite Score in the two groups and across time were adjusted for age, education and disease duration, using linear mixed models (LMM) with the Satterthwaite method, and multiple comparison outcomes were adjusted under the Bonferroni criterion. The group of A53T-PD or iPD and sex were used as fixed factors, participants as random effects, while age, years of education and disease duration as covariates. The same approach was adopted for the analysis regarding MDS-UPDRS.III.ON, H&Y.ON, MDS-UPDRS.IV, MDS-UPDRS.II, SCOPA-AUT, QUIP, S&E, but age, LEDD and disease duration were used as covariates. Finally, UPDRS.III.OFF, EPWORTH, RBDQ, UPDRS.IA, UPDRS.IA.1.1 (cognition), UPDRS.IA.1.2 (psychosis), UPDRS.Ib, GDS, and H&Y.OFF changes were also analyzed using LMM and age and disease duration were used as covariates. Missing follow-up values were handled by the LMM model. No subgroups were constituted, due to the low number of included participants. For the same reason, no sensitivity analysis was conducted.

The analysis was carried out using SPSS v 29.0 and the significance was set at 0.05 in all cases. Sample matching was done with MatchIt package v. 4.5 using R v4.3 as described in “participant selection” section, above.

## 3. RESULTS

### Participants

We have reviewed data regarding all genetic PD patients at enrollment (n=791), collected all SNCA A53T mutation carriers (n=42) and removed all GBA (n=303), LRRK2 (n=428), PRKN (n=14), PINK1 (n=1) and simultaneous GBA/LRRK2 (n=3) mutation carriers. From those collected we ruled out any prodromal-asymptomatic carriers of A53T mutation (n=11) to complete our first sample of SNCA A53T-PD participants (n=31). To acquire 3-year longitudinal data from this sample, we searched the number of patients who had completed baseline visit (BL) and Visit 8 (V8) of the PPMI Protocol, with secondary search criteria of Visit 4 (V4) and Visit 6 (V6) completion to be applied to the main criterion. V4 corresponds to year number 1, V6 corresponds to year number 2 and V8 corresponds to year number 3. We found 16 A53T-PD carriers who fulfilled both BL & V8 visits (study sample), while 13 and 11 of them had also completed V4 & V6 visits respectively.

Finally, 16 A53T-PD participants were eligible (13 A53T-PD from the center at the University of Athens and 3 from the center at the University of Salerno). As a result, 48 matched iPD participants were selected (3:1 ratio), as previously described.

Time points were given successive numbers 1, 2, 3 and 4, where they correspond to baseline, first year of follow-up, second year and third year respectively. Eight A53T-PD participants had 2 follow-up visits, instead of the full three, while eleven iPD participants had 2 follow-up visits.

### Demographic features at baseline

The demographic features at baseline are summarized in the **Table 1**. Age at baseline, duration of Parkinson’s disease (PD), sex and years of education were comparable in the two groups, although there were proportionally more men in iPD.

**Table 1.** Demographics features in A53T-PD vs iPD at Baseline.

| FEATURE | A53T-PD (N=16) | iPD (N=48) | p Value |
| --- | --- | --- | --- |
| *Demographics* |  |  |  |
|  | mean±SD | mean±SD |  |
| Age (years) | 50.938 (±11.862) | 52.919 (±9.474) | 0.598>0.05<sup>a,b</sup> |
| Disease duration (months) | 50.56 (±37.518) | 52.96 (±35.246) | 0.687>0.05<sup>a,b</sup> |
| Education (years) | 13.06 (±4.795) | 15.63 (±3.036) | 0.106>0.05<sup>a,b</sup> |
|  | frequencies | Frequencies |  |
| Sex (female / male) | 8 (50%) / 8 (50%) | 14 (29,2%) / 34 (70,8%) | 0.129>0.05<sup>a,c</sup> |

a: p-value=0.05,
b: Mann Whitney *U* test,
c: Chi-Square

### Medication

LEDD: a significant increase was observed in A53T-PD at times “2”, “3” and “4” vs time “1” (p=0.044, p=0.001 and p=0.001 respectively). A53T-PD had significantly higher values compared to iPD (p-values <0.001) at all time points.

### Nonmotor features

#### Cognitive Tests

Results are summarized in **Table 2** and **Figure 1**.

**Table 2.** Cognitive tests in A53T-PD vs iPD in a 3-Year Follow up.

| FEATURE |  | A53T-PD | lpd | p (between groups) |
| --- | --- | --- | --- | --- |
|  | Time | mean (±SD) | mean (±SD) |  |
| MoCA | 1 | 25.79 (±0.871) | 27.195 (±0.552) | 0.179 |
|  | 2 | 24.290 (± 1.01) | 27.803 (±0.635)<sup>[2][2]</sup> | 0.005 |
|  | 3 | 23.223 (±1.02)<sup>***1</sup> | 27.399 (±0.62)<sup>[2][2]</sup> | 0.001 |
|  | 4 | 21.635 (±1.223)<sup>***1**2</sup> | 27.659 (±0.771)<sup>[2][2]</sup> | <0.001 |
| Benton | 1 | 10.167 (±0.592) | 13.331 (± 0.370)<sup>+++</sup> | <0.001 |
|  | 2 | 9.872 (± 0.687) | 12.960 (±0.426)<sup>+++</sup> | 0.001 |
|  | 3 | 9.788 (± 0.786) | 13.004 (±0.458)<sup>+++</sup> | <0.001 |
|  | 4 | 8.497 (± 0.650)<sup>*1*2</sup> | 12.954 (±0.407)<sup>+++</sup> | <0.001 |
| LNST | 1 | 8.312 (±0.678) | 11.030 (± 0.425)<sup>+++</sup> | 0.001 |
|  | 2 | 7.919 (±0.915) | 11.490 (± 0.563)<sup>++</sup> | 0.002 |
|  | 3 | 7.348 (± 0.867) | 11.299 (± 0.481)<sup>+++</sup> | <0.001 |
|  | 4 | 6.410 (± 0.830)<sup>*1</sup> | 10.922 (± 0.521)<sup>+++</sup> | <0.001 |
| Phonemic F | 1 | 11.572 (±1.086) | 14.644 (±0.688)<sup>+</sup> | 0.02 |
|  | 2 | 8.352 (±1.288) | 15.145 (±0.795)<sup>+++</sup> | <0.001 |
|  | 3 | 10.240 (±1.476) | 16.119 (±0.815)<sup>+++</sup> | 0.001 |
|  | 4 | 8.506 (± 1.272)<sup>*1</sup> | 15.955 (±0.794)<sup>+++</sup> | <0.001 |
| SDMT | 1 | 31.220 (±2.974) | 44.497 (±1.858)<sup>+++</sup> | <0.001 |
|  | 2 | 24.699 (±3.113)<sup>***1</sup> | 43.551 (±1.897)<sup>+++</sup> | <0.001 |
|  | 3 | 20.608 (±3.171)<sup>***1</sup> | 44.798 (±1.839)<sup>+++</sup> | <0.001 |
|  | 4 | 23.245 (±3.740)<sup>***1</sup> | 46.694 (±2.308)<sup>+++</sup> | <0.001 |
| Semantic F | 1 | 18.592 (±1.703) | 22.044 (±1.073) | 0.092 |
|  | 2 | 16.455 (±1.705) | 22.513 (±1.059)<sup>††</sup> | 0.004 |
|  | 3 | 15.704 (±1.761) | 22.602 (±1.014)<sup>†††</sup> | 0.001 |
|  | 4 | 13.228 (±1.770)<sup>*1</sup> | 21.965 (±1.117)<sup>†††</sup> | <0.001 |
| HVLT-IM-REC | 1 | 20.452 (±1.548) | 26.976 (±0.968)<sup>†††</sup> | 0.001 |
|  | 2 | 19.844 (±1.884) | 25.101 (±1.164)<sup>†</sup> | 0.021 |
|  | 3 | 16.844 (±1.715) | 26.525 (±0.958)<sup>†††</sup> | <0.001 |
|  | 4 | 15.609 (±2.009)<sup>*1</sup> | 25.820 (±1.261)<sup>†††</sup> | <0.001 |
| HVLT-REC | 1 | 11.108 (±0.366) | 11.393 (±0.229) | 0.513 |
|  | 2 | 10.683 (±0.686) | 11.032 (±0.428) | 0.669 |
|  | 3 | 9.466 (±0.769) | 11.372 (±0.399)<sup>†</sup> | 0.033 |
|  | 4 | 9.512 (±0.808) | 10.967 (±0.509) | 0.134 |
| HVLT-RDLY | 1 | 7.202 (±0.765) | 8.974 (±0.48) | 0.055 |
|  | 2 | 7.731 (±0.838) | 8.419 (±0.517) | 0.488 |
|  | 3 | 6.615 (±0.841) | 9.425 (±0.461)<sup>††</sup> | 0.005 |
|  | 4 | 5.536 (±0.802)<sup>*2</sup> | 8.938 (±0.504)<sup>†††</sup> | <0.001 |
| Composite score | 1 | 18.826 (1.072) | 23.635 (0.671)<sup>†††</sup> | <0.001 |
|  | 2 | 16.567 (1.146)<sup>**1</sup> | 23.655 (0.712)<sup>†††</sup> | <0.001 |
|  | 3 | 15.155(1.133)<sup>***1</sup> | 23.855 (0.68)<sup>†††</sup> | <0.001 |
|  | 4 | 14.51(1.29)<sup>***1</sup> | 24.014 (0.801)<sup>†††</sup> | <0.001 |

1= Baseline; 2= year number 1; 3= year number 2; 4= year number 3; HVLT-IM-REC = Hopkins Verbal Learning Test Immediate Recall; HVLT-RDLY = Hopkins Verbal Learning Test Delayed Recall; HVLT-REC = Hopkins Verbal Learning Test Delayed Recognition; LNST = Letter Number Sequencing Test; MoCA = Montreal Cognitive Assessment; SDMT = Symbol Digit Modalities Test; BENTON= Benton Judgement of line orientation test; SEMANTIC F= Semantic fluency; Phonemic F= Phonemic verbal fluency; Composite score = (MOCA+LNST+SDMT+BENTON+SEMANTIC F)/5
<sup>\*1</sup> Statistically significant differences between the annotated time and 1
<sup>\*2</sup> Statistically significant differences between the annotated time and time 2
\*\* p<0.01 \*\*\* p≤0.001
<sup>†</sup> Statistically significant differences between case and controls at the specified time point
<sup>†</sup> p<0.05 <sup>††</sup> p<0.01 <sup>†††</sup> p≤0.001
p<sub>3</sub>: p-value between groups

**Figure 1.**
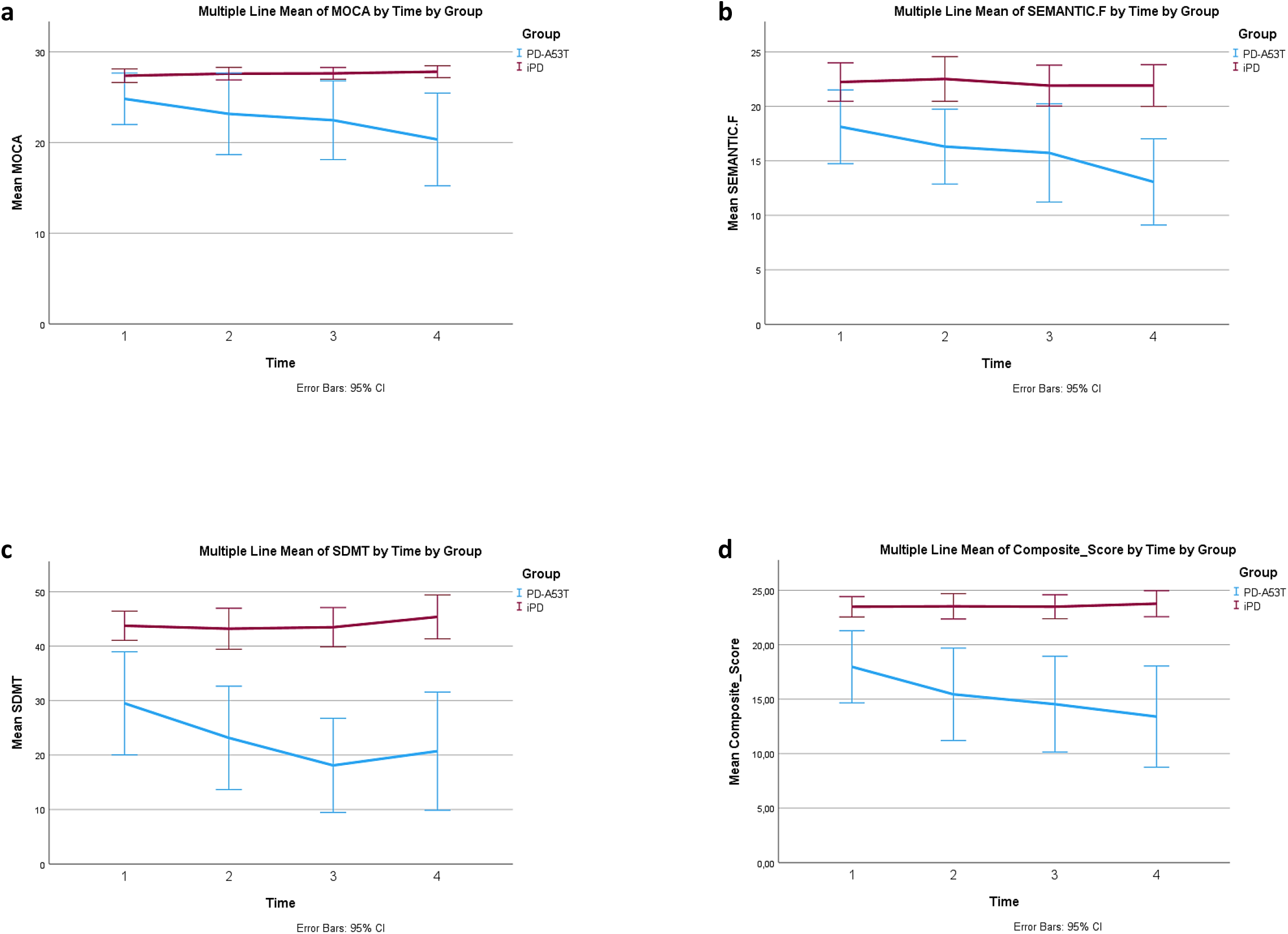
Longitudinal cognitive Tests in A53T-PD vs. iPD. **(a)** a significant reduction was observed in the MOCA score in the group of A53T-PD at time “3”(year number 2) vs time “1” (baseline), as well at time “4” (year number 3) comparing to times “1” (baseline) and “2” (year number 1). The two groups differed at all time points, except baseline, with p-values <0.01 indicating lower MOCA values in the group of A53T-PD. **(b)** A significant reduction in the SDMT score was observed in the group of A53T-PD at times “2”, “3” and”4” vs time “1”. The two groups differed at all time points, indicating lower SDMT values in the group of A53T-PD. **(c)** In Semantic Fluency, differences were found between groups but also across time for the group of A53T-PD. Specifically a significant reduction was observed in the group of A53T-PD at time “4” vs time “1”. The two groups differed at all time points, except baseline, indicating lower SEMANTIC F values in the group of A53T-PD. **(d)** A composite cognitive score was generated (COMPOSITE), constituted of 5 cognitive domains: MOCA score, Semantic fluency, Letter Number Sequencing Test, BENTON and Symbol Digit Modality Test [COMPOSITE= (MOCA + Semantic.F + LNST +BENTON +SDMT)/5]. A significant reduction was observed in the COMPOSITE score in the group of A53T-PD at time “2”(year number 1) vs time “1” (baseline), at time “3” (year number 2) vs time “1” (baseline), as well at “4” (year number 3) vs time “1”. The two groups differed at all time points, with p-values <0.001, indicating significant cognitive decline in the sum of these domains in the group of A53T-PD related to iPD.

##### MoCA score

A significant reduction was observed in A53T-PD at time “3” vs time “1”, and at time “4” compared to times “1” and “2”. A53T-PD had significantly lower values compared to iPD at all time points except baseline.

##### BENTON

A significant reduction was observed in A53T-PD at time “4” compared to times “1” and “2”. A53T-PD had significantly lower values compared to iPD at all time points.

##### LNST

A significant reduction was observed in A53T-PD at time “4” vs time “1”. A53T-PD had significantly lower values compared to iPD at all time points.

##### PHONEMIC.F

A significant reduction was observed in A53T-PD at time “4” vs time “1”. A53T-PD had significantly lower values compared to iPD at all time points.

##### SDMT

A significant reduction was observed in A53T-PD at times “2”, “3” and “4” vs time “1”. A53T-PD had significantly lower values compared to iPD at all time points.

##### SEMANTIC.F

A significant reduction was observed in A53T-PD at time “4” vs time “1”. A53T-PD had significantly lower values compared to iPD at all time points except baseline.

##### HVLT.IM-REC

A significant reduction was observed in A53T-PD at time “4” vs time “1”. A53T-PD had significantly lower values compared to iPD at all time points.

##### HVLT-REC

Differences were not found across time for either group. The two groups differed at time point “3”, indicating lower values in A53T-PD.

##### HVLT-RDLY

A significant reduction was observed in A53T-PD at time “4” vs time “2”. A53T-PD had significantly lower values compared to iPD at time points “3” and “4”.

##### Composite Score

A significant reduction was observed in A53T-PD at times “2”, “3” and “4” vs time “1”. A53T-PD had significantly lower values compared to iPD at all time points.

### Other non-motor features

Results are summarized in **Table 3** and **Figure 2**.

**Table 3.**
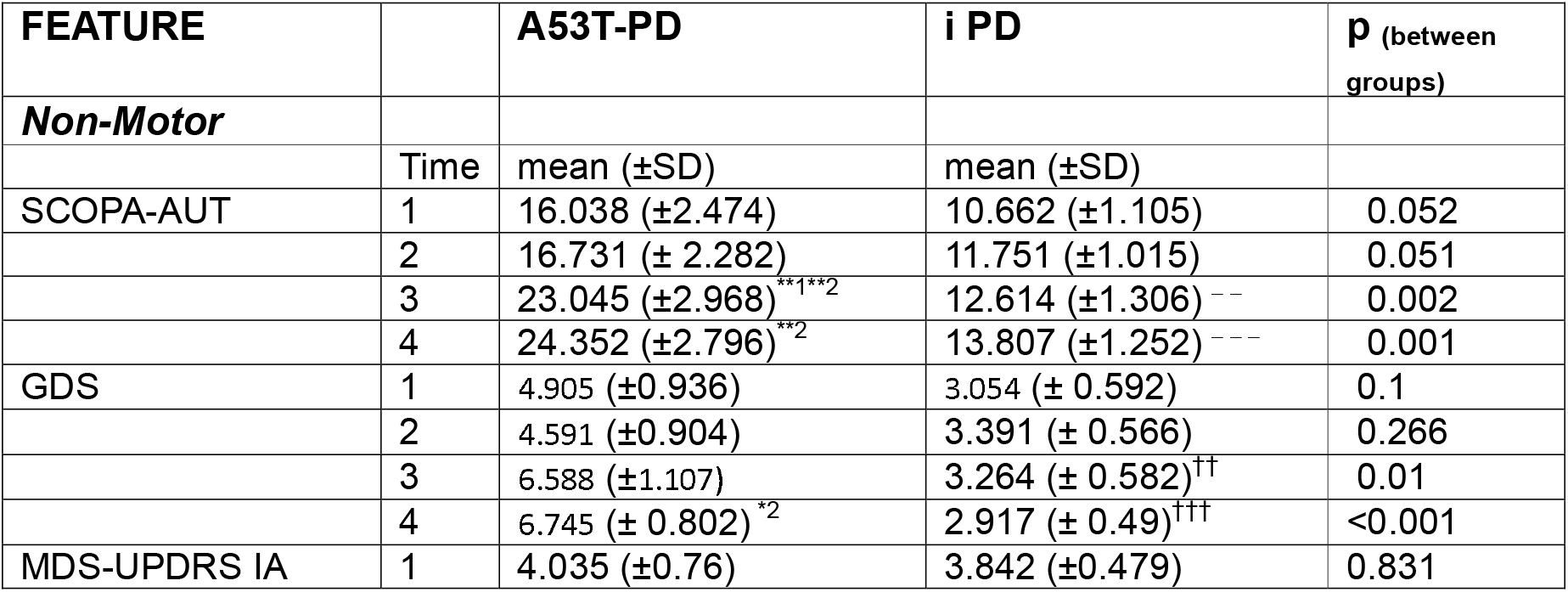
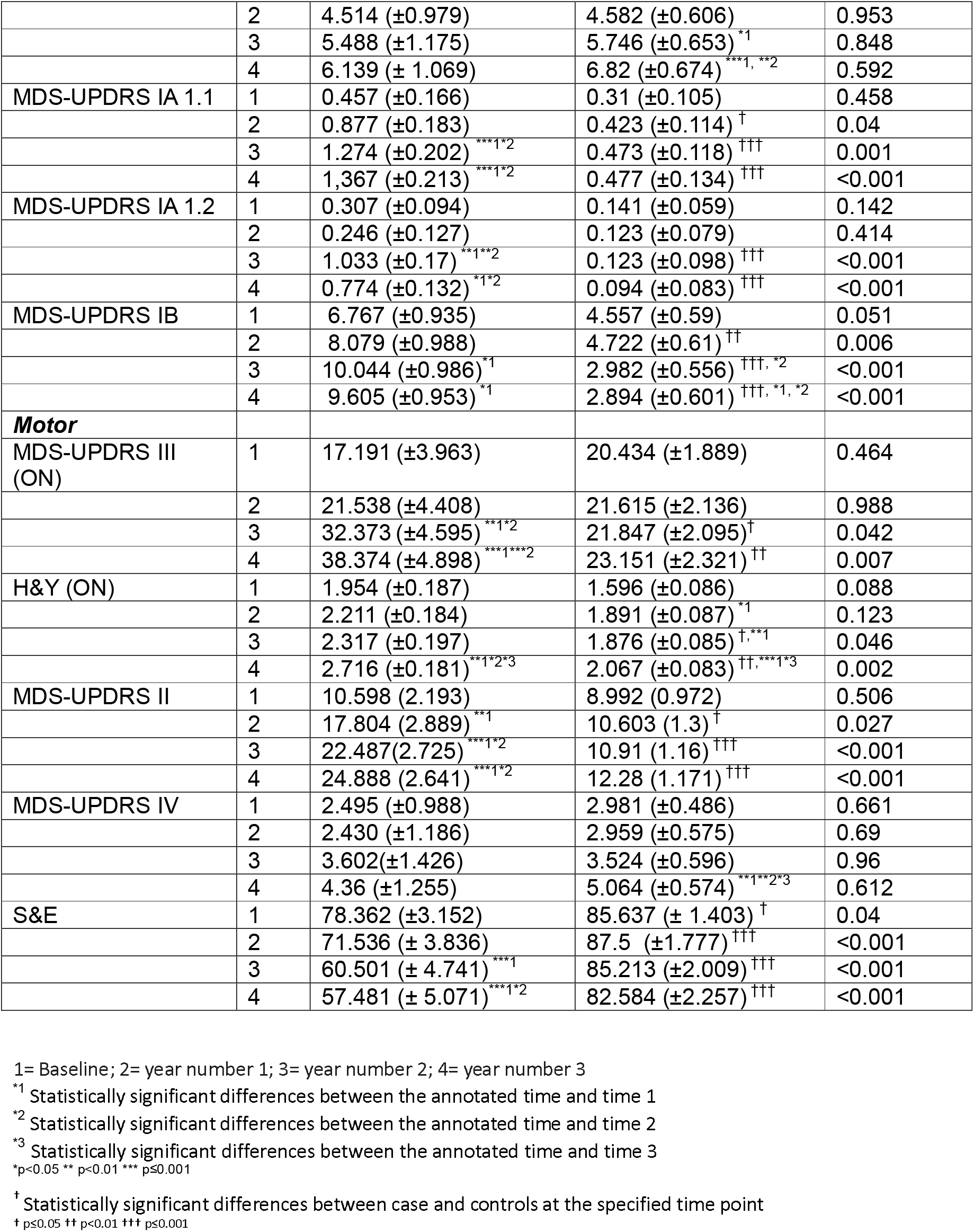
Other Motor and Non Motor features, beyond cognitive testing, in A53T-PD vs iPD in a 3-Year Follow up.

**Figure 2.**
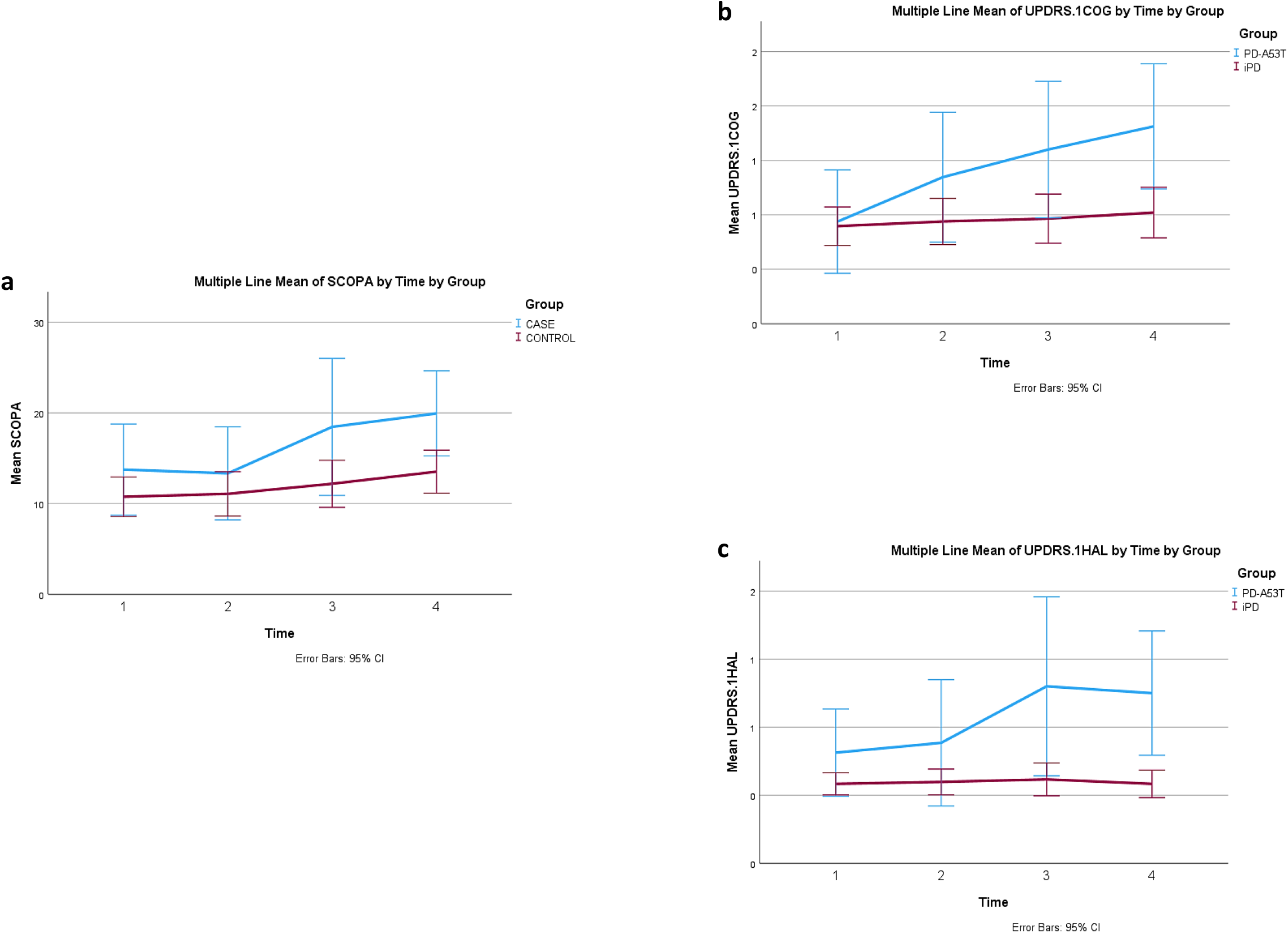
Longitudinal Non-Motor Tests in A53T-PD vs. iPD. **(a) SCOPA-AUT: a** significant increase was observed in the group of A53T-PD at times “3” vs time “1” and “2”, as well at time “4” comparing to time “2”. The two groups differed at time points “3” and “4”, indicating higher SCOPA-AUT values in the group of A53T-PD. **(b) MDS-UPDRS IA: 1.1 Cognition (UPDRS.1COG):** A significant increase was observed in the group of A53T-PD at times “3” and”4” vs time “1” and at times “3” and”4” vs time “2” The two groups differed at all time points except baseline, with p-values ≤0.04, indicating higher values in the group of A53T-PD. **(c) MDS-UPDRS IA: 1.2 Hallucinations and psychosis (UPDRS.1HAL): Cognition (UPDRS.1COG):** A significant increase was observed in the group of A53T-PD at times “3” and “4” vs time “1” and at times “3” and “4” vs time “2”. The two groups differed at time points “3” and “4”, with p-values <0.001, indicating higher values in the group of A53T-PD.

#### SCOPA-AUT

A significant increase was observed in A53T-PD at times “3” vs “1” and “2”, and at time “4” compared to time “2”. A53T-PD had significantly higher values compared to iPD at time points “3” and “4”.

#### MDS-UPDRS.IA

(Complex behaviors): A significant increase was observed in iPD at time “3” vs time “1”, and as at time “4” compared to times “1” and “2”. The two groups did not differ at any time point.

#### MDS-UPDRS.IA

1.1 Cognitive impairment: A significant increase was observed in A53T-PD at times “3” and “4” vs time “1” and at times “3” and “4” vs time “2”. A53T-PD had significantly higher values compared to iPD at all time points except baseline.

#### MDS-UPDRS.IA: 1.2 Hallucinations and psychosis

A significant increase was observed in A53T-PD at times “3” and “4” vs time “1”, and at times “3” and “4” vs time “2”. A53T-PD had significantly higher values compared to iPD at all time points “3” and “4”.

#### MDS-UPDRS.Ib

A significant increase was observed in A53T-PD at times “3” and “4” vs time “1” while a decrease was observed in the iPD group at time “4” compared to times “1” and “2” and at time “3” vs time “2”. A53T-PD had significantly higher values compared to iPD at all time points except baseline.

#### GDS

A significant increase was observed in A53T-PD differences were found across time in A53T-PD at time “4” vs time “2”. A53T-PD had significantly higher values compared to iPD at time points “3” and “4”.

In QUIP, RBDQ and EPWORTH there were no differences found between groups or across time for any group.

### Motor features

Results are summarized in **Table 3** and in **Figure 3**.

**Figure 3.**
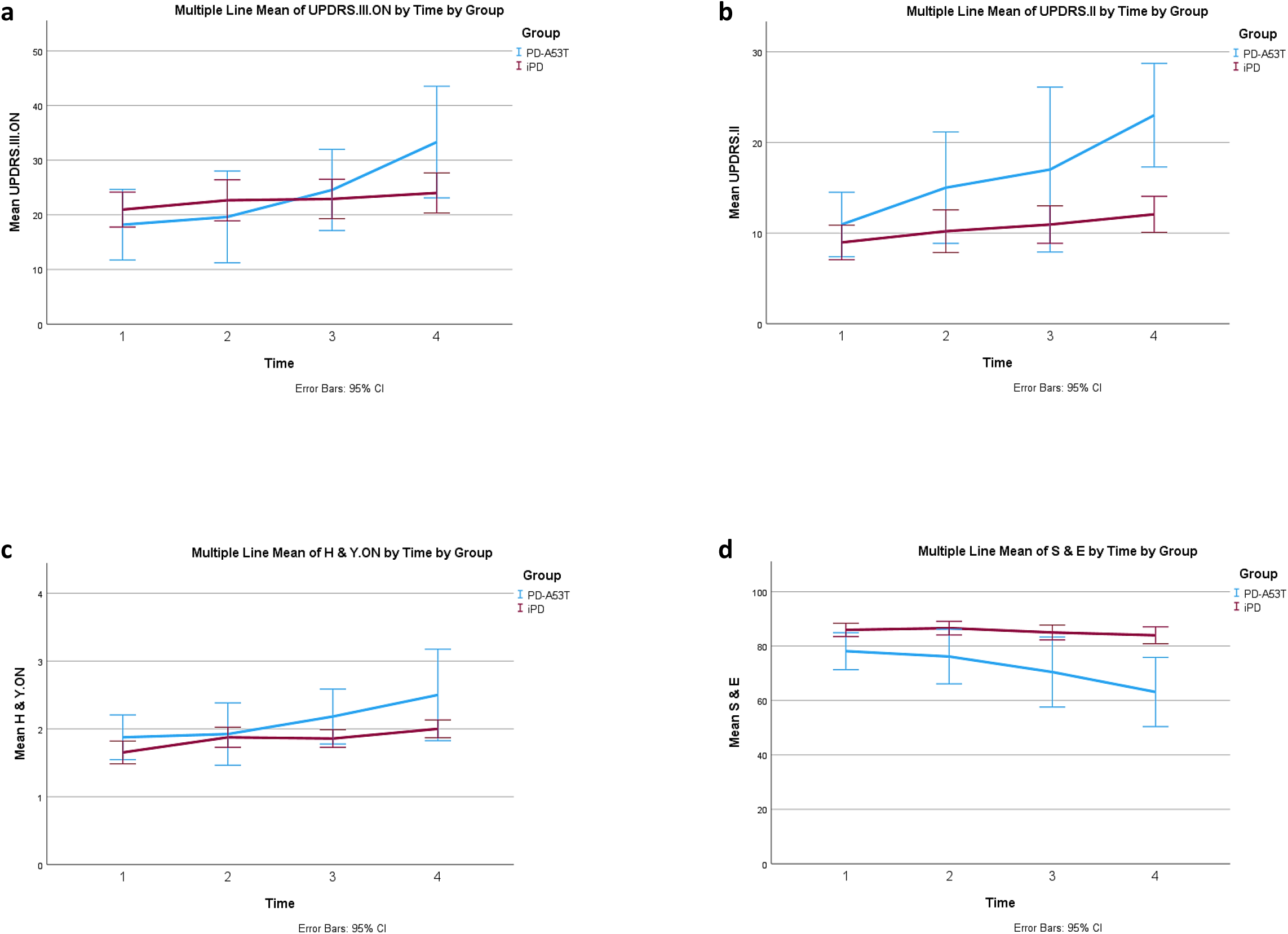
Longitudinal Motor Tests in A53T-PD vs. iPD. **(a) MDS-UPDRS-III in the ON state:** a significant increase was observed in the group of A53T-PD at times “3” vs “1” and “2”, as well at time “4” comparing to times “1” and “2”. The two groups differed at time points “3” and “4”, indicating higher UPDRS.III.ON values in the group of A53T-PD. **(b) H&Y scale in the ON state:** a significant increase was observed in A53T-PD at time “4” vs “1”, “2” and “3”. The two groups differed at time points “3” and “4”, indicating higher H & Y. ON values in the group of A53T-PD. **(c) MDS-UPDRS II:** For the group of A53T-PD all values were significantly different, gradually increasing, except for time “3” vs “4”. A statistically significant increase is observed between baseline and time “4” for the iPD group as well (p=0.005). The two groups differed at time points “2”, “3” and “4”, indicating higher UPDRS.II values in the group of A53T-PD. **(d) S&E:** a significant reduction was observed in the group of A53T-PD at time “3” vs time “1”, as well at time “4” comparing to times “1” and “2”. The two groups differed at all time points, indicating lower S &E values in the group of A53T-PD.

#### MDS-UPDRS.III.ON

A significant increase was observed in A53T-PD at times “3” vs “1” and “2”, as well at time “4” compared to times “1” and “2”. A53T-PD had significantly higher values compared to iPD at time points “3” and “4”.

#### H&Y. ON

A significant increase was observed in A53T-PD at time “4” vs “1”, “2” and “3”. Also, a significant increase was observed in iPD at time “4” vs time “1” and “3”, at time “3” vs time “1” and at time “2” vs time “1”. A53T-PD had significantly higher values compared to iPD at time points “3” and “4”.

#### S&E

A significant reduction was observed in A53T-PD at time “3” vs time “1”, as well at time “4” compared to times “1” and “2”. A53T-PD had significantly lower values compared to iPD at all time points.

#### MDS-UPDRS.II

A statistically significant increase was observed between baseline and time “4” in iPD. A significant increase was observed in A53T-PD at time “4” vs “1”, “2”, at time “3” vs “1” and “2” and at time “2” vs “1”. A53T-PD had significantly higher values compared to iPD at time points “2”, “3” and “4”.

#### MDS-UPDRS.IV

No differences were found between groups nor across time in A53T-PD. A significant increase was found across time only in iPD, at time “4” vs time “1”, “2”, and “3”.

#### H&Y.OFF and MDS-UPDRS.III.OFF

There were too many missing values in these assessments for them to be meaningfully assessed (data not shown).

## 4. DISCUSSION

The current work represents the most comprehensive, to date, longitudinal study of clinical features in p.A53T-PD compared to iPD. We compared 16 A53T-PD with 48 iPD patients, matched for age and disease duration at baseline, using several certified tests administered to these patients as part of their participation in the PPMI study. For the first time we report here on detailed assessments at multiple time points over a period of 3 years.

Detailed neuropsychological assessment revealed significant cognitive decline in A53T-PD compared to iPD, but also across time in A53T-PD. At baseline A53T-PD scored significantly lower in BENTON, LNST, PHONEMIC F, SDMT, and HVLT.IM-REC compared to iPD, indicating more impaired executive and visuospatial function. MOCA, Semantic F, HVLT-REC and HVLT-RDLY were also lower, but not significantly so. These findings are consistent with our previous study where we had compared cross-sectionally 18 p.A53T-PD with 18 matched iPD participating in the PPMI study^7^. Only 7 of the A53T-PD in our previous study^7^ are included herein. The consistency of results confirms a distinct cognitive profile in A53T-PD, reflecting a frontal-parietal network dysfunction at a juncture of about 4-5 years after motor disease onset.

Given the fact that MOCA, reflecting global cognitive function, was not significantly different at baseline between groups in the present study, or in our previous work^7^, with assessment of patients at similar time points of 4-5 years after disease onset, we decided to establish a composite score of multiple cognitive functions, as described in the “Methods” section. The cognitive Composite score reflects multiple domains of cognitive function, but is more powerful than MOCA, including 5 cognitive tests and emphasizing executive/visuospatial function, compatible with the pattern of cognitive impairment in PD. This score was significantly lower in A53T-PD at all time points, appearing to better reflect the cognitive decline within the early years of disease in A53T-PD. Another global measure of cognitive decline, the item 1.1 of MDS-UPDRS.IA, was also worse in A53T-PD at all time points and deteriorated significantly across time.

Importantly, the longitudinal nature of the study enabled us to establish that even cognitive measures that were not significantly different from iPD at baseline, worsened significantly in A53T-PD after 6-7 years of disease duration, while they remained stable in iPD. Not surprisingly, cognitive measures that were affected at baseline, such as the Composite Score, also continued to deteriorate in A53T-PD over time. This more widespread and severe cognitive dysfunction is likely related to the extensive burden of LB pathology, especially in cortical regions, that is known to occur in A53T-PD^9,10^.

Dysautonomic symptoms, as per the SCOPA-AUT scale, are more prominent over time in A53T-PD compared to iPD, unlike our previous cross-sectional study^7^. Statistically significant worsening of autonomic function was found also across time in A53T-PD, consistent with our previous study^6^. However, this needs to be validated with more objective measures. Using non-invasive autonomic function tests, we have preliminary evidence that A53T-PD perform worse compared to iPD or GBA-PD (Simitsi et al., unpublished results). These findings are in accordance with neuropathological studies in A53T-PD showing severe LB pathology in regions controlling autonomic function, such as the locus coeruleus and the Dorsal Motor Nucleus of the Vagus^10^.

As far as MDS-UPDRS.IA is concerned, we found that the two groups did not differ at any time point but, examining separate items, we found higher scores concerning cognition and psychosis (items 1.1 and 1.2 respectively) in A53T-PD compared to iPD, as well as a significant increase in the group of A53T-PD across time. These findings indicate that the use of more specific and targeted questionnaires and assessments may reveal a more distinct profile of the disease.

Scores in RBDQS were similar between the two groups, consistent with our previous findings^7^; given the poor predictive validity of such questionnaires for polysomnography-confirmed RBD^11^, especially in the absence of confirmation from a bed partner, these results should be taken with caution. In a previous study from our cohort of A53T-PD, a simple sleep history question in 22 participants indicated that 45% had symptoms suggestive of RBD^6^. In another study, assessing 15 p.A53T-carriers with simultaneous Video-PSG (polysomnography) recordings, we found that RBD occurs in the majority of PD-A53T (8/10), in contrast to most other genetic forms of PD. We also found a paucity of a sleep disorder in asymptomatic carriers, suggesting that they may have not yet reached the prodromal phase when such sleep disorders manifest^8^. Further PSG studies, in particular longitudinal, are needed in order to assess the evolution of sleep disorders in this rare group of participants.

At baseline A53T-PD performed similarly on motor function and examination compared to iPD, no doubt because the increased LEDD masked the more pronounced nigrostriatal degeneration detected with dopaminergic imaging ^12^. Despite an increase in the difference in LEDD between iPD and A53T-PD over time, A53T-PD eventually showed worse performance even on quite global motor assessments, such as H&Y. ON or S&E, but also on more detailed ones, such as MDS-UPDRS.II and MDS-UPDRS.III.ON. This again underscores the more severe progressive nature of motor dysfunction in this form of PD, merely hinted in our prior study^6^.

As far as MDS-UPDRS.IV is concerned, no differences were found between groups nor across time in A53T-PD, underscoring the fact that motor complications are not usually a main feature of advanced A53T-PD, in which a more global and continuous motor and non-motor dysfunction predominates.

Our study has some limitations, mainly the small number of participants in A53T-PD. The issue is compounded by the number of participants who performed visits at the one- and two-year time points, which is even lower, accounting in part for the larger variability within this group. However, the differences observed across groups and across time are generally of large magnitude and of high statistical significance; there is consistency between the current and prior studies from our group in various measures, while the novel findings of accelerated disease progression are reflected in a variety of different examination modalities and are internally consistent. The fact that iPD patients show little deterioration over 3 years is probably due to their relatively young age, matched to the age of A53T-PD. The generalization of our results is difficult, due to variable genetic background. While PPMI includes persons from a wide ethnic and genetic background, A53T-PD are predominantly Caucasians from the Mediterranean basin.

The overall picture of p.A53T carriers, and especially the current longitudinal data, indicate that this genetic variant represents a more severe disease form compared to iPD, with accelerated decline in various motor and non-motor measures. Cognitive dysfunction is especially interesting in this regard, as it is not easily modified by current drug interventions. We suggest that such parameters could be used as endpoints in clinical trials with disease-modifying agents. Of special interest are agents that would counteract the pathological effects of AS, as this cohort represents a unique case scenario in which, undoubtedly, disease etiopathogenesis is directly related to pathological AS. This provides the basis of the idea to perform in this population proof-of concept clinical trials with disease-modifying agents targeting AS. LB pathology is ubiquitous in almost all cases with iPD, something brought to the forefront with the advent of the highly sensitive and specific AS seeding amplification Assays (SAAs)^13^. Therefore, combined genetic and neuropathological data indicate that the SNCA gene and the AS protein are highly relevant for iPD and represent valid therapeutic targets. Hence, insights gained from the study of this rare genetic synucleinopathy may impact globally the understanding and care of the vast majority of patients with iPD.

## Data Availability

The data used in the preparation of this article were obtained [on October 05, 2023] from the Parkinson's Progression Markers Initiative (PPMI) database (https://www.ppmi-info.org/access-data specimens/download-data), RRID:SCR_006431. For up-to-date information on the study, visit http://www.ppmi-info.org. This analysis used data openly available from PPMI.

## Acknowledgements

PPMI - a public-private partnership - is funded by the Michael J. Fox Foundation for Parkinson’s Research and funding partners, including 4D Pharma, Abbvie, AcureX, Allergan, Amathus Therapeutics, Aligning Science Across Parkinson’s, AskBio, Avid Radiopharmaceuticals, BIAL, BioArctic, Biogen, Biohaven, BioLegend, BlueRock Therapeutics, Bristol-Myers Squibb, Calico Labs, Capsida Biotherapeutics, Celgene, Cerevel Therapeutics, Coave Therapeutics, DaCapo Brainscience, Denali, Edmond J. Safra Foundation, Eli Lilly, Gain Therapeutics, GE HealthCare, Genentech, GSK, Golub Capital, Handl Therapeutics, Insitro, Jazz Pharmaceuticals, Johnson & Johnson Innovative Medicine, Lundbeck, Merck, Meso Scale Discovery, Mission Therapeutics, Neurocrine Biosciences, Neuron23, Neuropore, Pfizer, Piramal, Prevail Therapeutics, Roche, Sanofi, Servier, Sun Pharma Advanced Research Company, Takeda, Teva, UCB, Vanqua Bio, Verily, Voyager Therapeutics, the Weston Family Foundation and Yumanity Therapeutics.

Additional support for this work was provided by the National Network for Research of Neurodegenerative Diseases on the basis of Medical Precision (Grant 2018 E01300001), funded by the General Secretariat of Research and Innovation (GSRI), and by Brain Precision (TAEDR-0535850), funded by the GSRI, through funds provided by the European Union (Next Generation EU) to the National Recovery and Resilience Plan.

